# Automatic sleep staging in patients with suspected sleep disorders: a comparison of existing methods on portable setups

**DOI:** 10.64898/2026.07.06.26357378

**Authors:** Katarina M. Gunter, Alexis Dorier, Francesca Bowring, Gary Dennis, Christine Lo, Timothy Quinnell, Mkael Symmonds, Pietro-Luca Ratti, Michele T. Hu, Mauricio Villarroel

## Abstract

**Background:** Automatic sleep staging algorithms are increasingly applied in clinical and home-based recordings. However, their performance may degrade when transferred to new montages and clinical populations. This is particularly relevant in reduced-channel portable PSG and in disorders such as REM sleep behaviour disorder (RBD), where altered sleep architecture may challenge pretrained models.

**Objective:** To evaluate and compare multiple open-source sleep staging algorithms on a minimal portable PSG setup in controls and patients with and without RBD, and to assess the impact of fine-tuning on clinic-ascertained data.

**Methods:** Six open-source models were applied to 76 subjects recruited from three clinical sleep medicine sites. Performance was assessed using accuracy, F1 scores, and Cohen’s, both overall and per sleep stage. Each model was evaluated out-of-the-box and after fine-tuning on clinical data.

**Results:** Out-of-the-box performance varied substantially across models (*κ* 0.21–0.54). Fine-tuning consistently improved agreement, with the best-performing model (GSSC) reaching *κ* = 0.58 indicating moderate to good agreement. Performance was highest in controls and lower in patient groups. N3 was the most reliably classified stage across models, whereas N1 remained consistently challenging. REM classification improved after fine-tuning in several architectures but remained model, and subgroup-dependent, particularly in RBD subjects.

**Conclusion:** Fine-tuning substantially mitigates domain shift, updating model parameters to align with new data distributions, when applying automatic sleep staging algorithms to portable clinical recordings. Model architecture influences robustness, with feature-learning approaches demonstrating greater adaptability than fixed-feature models. Despite moderate agreement after adaptation, performance, especially for REM and N1 remains insufficient for fully automated diagnostic use in clinical populations.

## 1 Introduction

Manual sleep staging, first standardised by Rechtschaffen and Kales in 1968 and later refined by the American Academy of Sleep Medicine (AASM), remains a routine practice in sleep clinics worldwide [1, 2]. The process involves classifying 30-second intervals, or “epochs,” of electrophysiological signals recorded during overnight video polysomnography (vPSG) into five stages: wake, non-REM 1 (N1), non-REM 2 (N2), non-REM 3 (N3), and rapid eye movement (REM) sleep. Beyond providing insight into the physiological mechanisms of sleep, sleep staging is also essential for the clinical diagnosis of sleep disorders. The gold standard for sleep staging, and subsequent diagnosis, is an in-clinic video polysomnography (vPSG) in which electrophysiological signals are measured by electroencephalography (EEG), electrooculography (EOG), electromyography (EMG), electrocardiogram (ECG), as well as various signals of respiratory effort, and video [2, 3]. However, in-clinic vPSG has several limitations, including restricted access due to limited bed capacity and specialised resources, as well as the need for referral to dedicated sleep centers. Patients often report poorer sleep quality in the hospital environment, which can in turn influence the validity of the study [4]. This has lead to greater interest in at-home based PSG set-ups. Home PSG is typically only used when obstructive sleep apnea (OSA) is suspected, whereas other sleep disorders still require a full PSG montage [5].

Manual sleep staging is highly resource-intensive, requiring trained experts to score an entire night of sleep, 30 seconds at a time. Automatic sleep staging has been a topic of research since the original standardisation of the procedure. Multiple open-source algorithms have been developed, from classical machine learning such as random forests, to deep learning using convolutional, recurrent neural networks (CCNs, RNNs), and more recently transformers. Reliable automatic sleep staging also has the potential to greatly improve the accessibility of sleep analysis and diagnosis. However, the majority of benchmark algorithms are trained and validated on the full in-clinic PSG montage. Whilst there has been a focus on reduced montage sleep staging in recent years, these models have been evaluated largely in healthy controls. There is a limited understanding of the robustness of previously published automatic sleep staging algorithms on portable at-home setups, and how these perform in a clinical cohort in patients with sleep disorders. This is particularly relevant in the context of REM-sleep-behaviour disorder (RBD). RBD is a parasomnia, with the main clinical symptom being dream enactment behaviour during REM due to REM sleep without atonia (RSWA) [6]. The accurate detection of REM sleep in this population is therefore challenging, particularly for automatic algorithms. Isolated RBD (iRBD) is also a prodromal state to alpha-synucleinopathy, with approximately 80% of patients going on to develop Parkinson’s disease (PD), dementia with lewy bodies (DLB), or multiple system atrophy (MSA) within 12 years from RBD diagnosis [7]. Therefore, beyond the burden to patients and bed-partners, early diagnosis of iRBD is also important for enabling timely risk stratification, patient monitoring, and recruitment into targeted neuroprotective clinical trials at an earlier stage when disease-modifying interventions may be most effective.

To our knowledge, the comparison of sleep staging algorithms in a clinical population is still a widely understudied topic. Previous works have examined the effect of training and evaluating models on different datasets, or the amount of training data and variability [8, 9, 10]. A recent study compared the performance of five sleep staging algorithms on a cohort of patients with chronic insomnia, and found variable performance across all sleep stages [11]. More recently, large-scale studies have begun to investigate the generalisability of sleep staging models across heterogeneous datasets and acquisition settings. For example, a foundation model approach demonstrated that performance is highly sensitive to dataset composition, channel availability, and cohort differences, highlighting persistent challenges in cross-cohort transfer and real-world deployment [12]. Recent work has also highlighted the lack of standardised and fair benchmarking frameworks for sleep staging algorithms. A large-scale evaluation across diverse datasets and acquisition settings demonstrated substantial variability in model performance and persistent demographic and clinical biases, with no single architecture consistently generalising across cohorts [13]. Studies using portable PSG devices are similarly limited, previous studies focus on device validation through the comparison of a given automatic staging algorithm versus manual staging based on the in-clinic PSG [14, 15]. To date, there has only been one previous publication comparing automatic sleep staging models in patients with RBD [16]. This study compared two models on PSG data collected in-clinic in a group of RBD patients compared to sex and age-matched patients with obstructive sleep apnea (OSA). There remains an unmet need for external validation of sleep-staging algorithms to assess their generalisability and performance in real-world clinical settings, as well as their potential integration into portable systems that could enhance accessibility and reduce time to diagnosis.

The primary objective of this work was to evaluate whether open-source automatic sleep staging algorithms can reliably stage sleep in patients referred to a sleep clinic for suspected sleep disorders, when applied to a portable, reduced-montage PSG setup. We systematically evaluated six open-source automatic sleep staging algorithms which vary in terms of architecture, input modalities, number of channels, and the type and source of the training data (Random forest classifier [17], YASA [18], GSSC [19], DeepSleep [10], SleepTransformer [20], and AttnSleep [21]). We performed the evaluation using a portable, reduced montage PSG setup recorded in clinic in parallel with the gold-standard vPSG, in a cohort of patients referred to a sleep clinic for suspected sleep disorders. Automatic sleep staging was compared to manual ground truth annotations by experts. The benchmark algorithms were first evaluated in their out-of-the-box form and subsequently fine-tuned or re-trained on a subset of clinical data from our cohort. Model performance was assessed based on agreement with manual scoring, as well as generalisation to our dataset and the potential clinical utility of these models for sleep staging. As a secondary analysis, we compare sleep staging in subjects with RBD versus non-RBD.

## 2 Materials and methods

### 2.1 Dataset

The dataset used in this work is part of a clinical study conducted at three sleep medicine sites; John Radcliffe Hospital (Oxford, UK), Royal Hallamshire Hospital (Sheffield, UK), and Royal Papworth Hospital (Cambridge, UK). This project is a collaboration between the Nuffield Department of Clinical Neurosciences (NDCN) and the University of Oxford, supported by funding from the National Institute for Health Research (NIHR) Oxford Biomedical Research Centre (BRC). It was approved by the South Central – Oxford Research Ethics Committee with the reference number 17/SC/0631. The primary objective of the study is to develop novel technologies for the remote diagnosis and monitoring of sleep disorders, primarily RBD.

#### 2.1.1 Study design

Participants were referred for vPSG assessment by their physician for suspected sleep disorders. They were invited to participate in the study during their vPSG assessment. Participants were required to be aged 18 years or older, with no evidence of significant cognitive impairment that could have limited their ability to provide informed consent. They could not present any medical or psychiatric condition that might have interfered with completion of the study assessments. Participants were excluded if they were pregnant, actively enrolled in a drug trial, or taking medication judged by the Principal Investigator to significantly affect sleep or recording quality. They were also excluded if they were actively using Continuous Positive Airway Pressure (CPAP) therapy or Deep Brain Stimulation. Furthermore, participants could not exhibit sensitivity to the electrode adhesive used for the EEG, ECG, and EMG sensors. Control participants were required to be in good health, with no prior diagnosis of RBD or PD, and no self-reported sleep problems.

Following verbal and written consent, a clinical evaluation was conducted by a member of the study team. This evaluation included an interview covering the participant’s medical history, past and current medications, exposure to alcohol, smoking and caffeine, and family history of sleep or neurodegenerative disorders. The interview was followed by a clinical examination assessing potential motor signs of parkinsonism, olfactory function, and cognitive performance through the Montreal Cognitive Assessment (MoCA) [22] and Informant Questionnaire on Cognitive Decline in the Elderly (IQCODE) [23]. Cognitive and motor function were further assessed using the Hoehn and Yahr scale [24], the Purdue Peg Board test [25] to evaluate manual dexterity, and an apathy assessment. Physiological parameters, including height, weight, and blood pressure (measured in both supine and standing positions), were also recorded.

Participants meeting the inclusion criteria after the clinical assessment underwent an overnight vPSG at the in-hospital sleep clinic under the supervision of a sleep technician. In addition to the gold-standard vPSG setup, participants were fitted with portable vPSG devices, including wrist actigraphy, fingertip Photoplethysmography (PPG), Electromyography (EMG), Electrocardiography (ECG), Electroencephalography (EEG), Electrooculography (EOG) electrodes, and a respiratory belt. Both the clinical gold-standard vPSG setup and the portable system recorded the same night of sleep in parallel.

After the vPSG recording, a sleep clinician scored the sleep–wake stages according to the American Academy of Sleep Medicine (AASM) guidelines [26] in 30-second epochs. The clinician evaluated the presence of sleep disorders such as RBD, obstructive sleep apnoea, or central sleep apnoea.

#### 2.1.2 Instrumentation

##### Clinical gold-standard polysomnography setups

The three recording sites used broadly similar vPSG setups, with site-specific differences in devices and channel configurations as detailed in Table 1. At the John Radcliffe (JR) hospital and Sheffield hospital, data were collected using Natus EMU40 or Natus Trex devices (Natus, Wisconsin, USA). The JR setup included a 20-channel Electroencephalography (EEG) montage, a 2-channel Electrooculography (EOG) configuration (left and right), Electromyography (EMG) from two chin electrodes and bilateral anterior tibialis muscles, and a 2-lead Electrocardiography (ECG), all sampled at 200 Hz or higher. Additional physiological signals comprised pulse rate and peripheral oxygen saturation (SpO_2_) from a pulse oximeter, airflow measured via nasal cannula and thermistor, respiratory effort assessed using abdominal and thoracic plethysmography belts, and body position derived from a 3-axis accelerometer, all recorded at the same sampling rate. The Sheffield setup included EEG with a 20-channel montage, EOG, EMG, ECG, pulse oximetry (pulse rate and SpO_2_), airflow measured via nasal cannula, respiratory effort recorded using abdominal and thoracic plethysmography belts, and body position derived from a 3-axis accelerometer. All signals were sampled at 200 Hz or higher.

At the Papworth site, data were recorded using the Natus Embla device. The EEG configuration consisted of four channels (two central and two occipital), while EOG (left and right) and EMG (three chin electrodes and bilateral anterior tibialis muscles) were sampled at 200 Hz. Pulse rate and SpO_2_ were captured using a pulse oximeter at 3 Hz. Airflow was measured via nasal cannula and thermistor at 20 Hz, and respiratory effort was assessed with abdominal and thoracic plethysmography belts at 10 Hz. Body position was derived from a 3-axis accelerometer and recorded at 10 Hz.

**Table 1:** Summary of the video polysomnography (vPSG) setup across the three recording sites. * At JR and Sheffield, all channels are stored at a uniform system rate of 200 Hz at a minimum. Derived oximetry values (SpO_2_, pulse rate) have a substantially lower effective update rate.

| Site | Device | Signal | Description | Sampling frequency [Hz] |
| --- | --- | --- | --- | --- |
| **JR and Sheffield** | Natus EMU40/Natus Trex | EEG | 20-channel montage | 200 |
|  |  | EOG | 2-channel (left & right) | 200 |
|  |  | EMG | 2 chin, 2 bilateral anterior tibialis | 200 |
|  |  | ECG | 2-lead | 200 |
|  |  | Pulse Rate | Pulse oximeter | 200* |
|  |  | SpO<sub>2</sub> | Pulse oximeter | 200* |
|  |  | Air Flow | Nasal cannula | 200 |
|  |  | Respiratory Effort | Abdominal and thoracic plethysmography belt | 200 |
|  |  | Body Position | Derived from 3-axis accelerometer | 200 |
| **Papworth** | Natus Embla | EEG | 2 central & 2 occipital | 200 |
|  |  | EOG | 2-channel (left & right) | 200 |
|  |  | EMG | 3 chin, 2 bilateral anterior tibialis | 200 |
|  |  | Pulse Rate | Pulse oximeter | 3 |
|  |  | SpO<sub>2</sub> | Pulse oximeter | 3 |
|  |  | Air Flow | Nasal cannula & thermistor | 20 |
|  |  | Respiratory Effort | Abdominal and thoracic plethysmography belt | 10 |
|  |  | Body Position | Derived from 3-axis accelerometer | 10 |

##### Portable video polysomnography setup

The SOMNOtouch RESP vPSG device (SOMNOmedics, Germany) was worn simultaneously with the clinical gold-standard vPSG setup in hospital to obtain parallel physiological and electrophysiological recordings. It provided measurements of body position, activity, oxygen saturation, heart rate, respiratory effort, airflow, and electrophysiological signals, as detailed in Table 2. Body position was inferred from tri-axial accelerometry placed on the participant’s chest and classified into five categories: upright, supine, right, left or prone, sampled at 1 Hz. Activity was measured using a tri-axial accelerometer worn on the dominant wrist and expressed as the magnitude of the combined x, y, and z axes, sampled at 16 Hz. SpO_2_ and pulse rate were obtained from a pulse oximeter and recorded at 4 Hz. Respiratory rate was derived from the piezoresistive thoracic and abdominal belts, facilitating the differentiation between central and obstructive sleep apnea, with signals sampled at 32 Hz. Airflow was measured using a nasal cannula at a sampling rate of 256 Hz. Electrophysiological recordings included a 1-lead ECG, a 1-channel EEG (right side frontal electrode referenced to contralateral mastoid), and a 2-channel EOG, all sampled at 256 Hz. EMG was recorded from one anterior tibialis muscle and one submentalis chin electrode, sampled at 64 Hz.

We applied quality control checks to each 30 second epoch from the EEG, EOG, and EMG channels recorded by the portable vPSG set up. These channels were specifically evaluated because they form the input signals for the open-source sleep staging algorithms used in this work. First, flat signal segments were identified by computing the sample standard deviation within each 30-second epoch for each EEG, EOG, and EMG channel. Epochs with a standard deviation of zero were excluded, as these indicated non-varying signal segments. Second, a clipping check identified signal saturation by rejecting epochs in which more than 5% of samples were equal to the minimum or maximum recorded values. Third, a quantisation check was performed to detect low-resolution or discretised signals; epochs containing fewer than 20 unique values were excluded. An epoch was considered valid only if it passed all three criteria. Channel-level signal quality was quantified as the proportion of valid epochs within each channel. An overall recording quality score was computed as the mean of all channel-level scores. Channels with a quality score below 0.5 were classified as poor quality and flagged for exclusion.

**Table 2:** Summary of the physiological signals and vital sign recorded by the SOMNOtouch RESP.

| Signal | Description | Sampling frequency [Hz] |
| --- | --- | --- |
| Body position | 3-axis accelerometer placed on the subject’s chest: upright, supine, left, right, or prone | 1 |
| Activity | 3-axis accelerometer placed on the subject’s wrist ($x, y, z$) | 16 |
| SpO<sub>2</sub> | Peripheral oxygen saturation computed using a pulse oximeter | 4 |
| HR | Heart rate computed using a pulse oximeter | 4 |
| Respiratory rate | Computed from a thoracic and an abdominal belt | 32 |
| Air flow | Computed using a nasal cannula | 256 |
| ECG | 1-lead electrocardiogram | 256 |
| EEG | 3-channels electroencephalogram | 256 |
| EOG | 2-channels electroculogram | 256 |
| EMG | 1 tibialis channel and 1 submental channel | 64 |

#### 2.1.3 Dataset description

A total of 102 participants were recruited, comprising 84 “Patient” participants referred for vPSG screening and 18 “Control” participants. Data acquisition was successful in the majority of the recording sessions. However, occasional software malfunctions or sensor coupling issues led to the loss or corruption of signals from clinical gold-standard and portable vPSG devices. A total of 76 recordings were acquired where both systems were used in parallel with available sleep stage annotations. The data quality checks described in Section 2.1.2 resulted in the exclusion of a further 17 recordings due to signal quality failure in the portable vPSG recordings. Table 3 summarises the resulting dataset across the three sites. The dataset was comprised of healthy control subjects, patients with RBD, and patients without RBD but diagnosed with a different sleep disorder (non-RBD patients).

### 2.2 Algorithm implementation

The algorithms compared in this work (Random forest classifier [17], YASA [18], GSSC [19], DeepSleep [10], SleepTransformer [20], and AttnSleep [21]) vary in terms of architecture, input modalities, number of channels, and the type and source of the training data (see Table 4. We have included a wide range of models, from classical tree-based methods to novel deep learning architectures. These models were trained on varying amounts of data from healthy controls, mixed cohorts, or specific sleep disorders.

**Table 3:** Summary of the subject groups and data recorded among the three recording sites. Age is reported as mean *±* SD (in years) per gender within each group. The *“of which RBD”* sub-row reports the subset of patients with confirmed RBD.

| Site | Participant group | No. of participants | Male (n) | Male age | Female (n) | Female age |
| --- | --- | --- | --- | --- | --- | --- |
| Sheffield hospital | *Patient* | 13 | 10 | 50.6 ± 16.3 | 3 | 54.0 ± 24.2 |
| | *of which RBD* | 5 | 5 | 61.4 ± 7.1 | 0 | — |
|  | *Control* | 0 | 0 | — | 0 | — |
| Papworth hospital | *Patient* | 13 | 7 | 57.6 ± 9.5 | 6 | 42.8 ± 12.8 |
| | *of which RBD* | 5 | 5 | 60.0 ± 8.1 | 0 | — |
| | *Control* | 3 | 0 | — | 3 | 76.3 ± 11.4 |
| JR hospital | *Patient* | 20 | 15 | 60.9 ± 16.1 | 5 | 49.4 ± 19.6 |
| | *of which RBD* | 10 | 8 | 68.2 ± 4.6 | 2 | 69.5 ± 6.4 |
| | *Control* | 10 | 8 | 75.4 ± 8.3 | 2 | 82.0 ± 7.1 |
| **Total per group** | *Patient* | 46 | 32 | 57.0 ± 15.2 | 14 | 47.6 ± 17.1 |
| | *of which RBD* | 20 | 18 | 64.1 ± 7.1 | 2 | 69.5 ± 6.4 |
| | *Control* | 13 | 8 | 75.4 ± 8.3 | 5 | 78.6 ± 9.3 |
| **Total** |  | **59** | **40** | **60.7 ± 15.9** | **19** | **55.7 ± 20.7** |

#### 2.2.1 YASA

Yet Another Sleep Algorithm (YASA) is a gradient-boosted decision tree ensemble (LightGBM) sleep staging classifier which utilises one EEG channel, as well as an EMG and EOG channel as inputs [18]. Time-domain features are extracted from each of the modalities, as well as spectral features from the EEG, for each 30-second epoch. The model was trained on 3,163 overnight PSG recordings from seven datasets, encompassing healthy participants and individuals with sleep disordered breathing or insomnia, with ages 6–89 years. The YASA algorithm takes into account temporal context through the use of a moving window, such that features from previous and future epochs contribute to classifying the current epoch. The authors compared the performance of YASA to two previously published sleep staging algorithms on two external datasets, and found that YASA had a similar performance in both healthy subjects and those with obstructive sleep apnoea.

#### 2.2.2 Random forest for sleep staging in RBD

The random forest sleep staging classifier developed by Cooray et al. in 2019 [17], was the first multi-stage classifier specifically for patients with RBD. The sleep staging model being the first block in a fully automated pipeline for the detection of RBD. The model was trained on diagnositc PSG’s from 53 patients with RBD, and 53 healthy controls. This model used three input channels: EEG (C4-A1 or C3-A2), the EOG (delta of the right and left channel), and the chin EMG (chin - submentalis). The model uses well known time- and frequency-domain features, extracted for each 10-second window from each of the modalities, and averaged over a 30-second epoch. The authors found that, in general, the model had poor performance in subjects with RBD compared to controls, particularly for REM staging, but that the performance was still within moderate agreement with the human expert.

#### 2.2.3 GSSC

The Greifswald Sleep Stage Classifier (GSSC) is a sleep-staging algorithm that combines convolutional and recurrent neural network architectures [19]. It uses a 1-dimensional ResNet-style convolutional network to extract features from both EEG and EOG channels. These channels are processed separately, allowing the model to operate flexibly in dual-channel mode (EEG + EOG) or single-channel mode (EEG only or EOG only). Each 30-second signal segment is converted into a compact feature vector, which is then passed to a classification block that can run in two modes: context-free and context aware. The context-free mode simply maps the feature vector to a sleep stage, whereas the context aware mode uses a gated recurrent unit (GRU) to incorporate information from past and future epochs into the classification of the current epoch. The authors compared the GSSC to other widely used sleep-staging algorithms and found that while its overall performance was comparable to other state-of-the-art models, the GSSC exhibited lower variance across different EEG channels, highlighting its robustness.

#### 2.2.4 DeepSleep

The DeepSleep algorithm [10] was focused on the effect of using multi-cohort data to improve out-of-sample sleep stage predictions, resulting in the largest training and validation dataset (15,684 PSG recordings) out of the models considered in this work. Five datasets were included in the analysis, from multiple cohorts, including PSG recordings from both healthy controls and patients with sleep disorders (mainly sleep disordered breathing). DeepSleep is a 1-D convolutional ResNet, that uses a minimal subset of the full PSG montage as input - specifically one central EEG, both the left and right EOG, and single submentalis (chin) EMG. A bidirectional GRU adds temporal context from neighboring epochs. DeepSleep’s performance improved with more training data, especially when the data spanned diverse cohorts.

#### 2.2.5 SleepTransformer

The SleepTransformer algorithm [20] predicts sleep stages based on the spectral representation of the input signal. The model uses a single EEG channel as input (Fpz-Cz or C4-A1), which is transformed into a time-frequency image using the short-time-fourier transformer (STFT), which is then fed into a two-level transformer. The spectrogram from one 30-second epoch is first processed by the epoch-transformer, outputting an epoch vector in which each window is weighted by attention scores. A window containing L consecutive epoch vectors are then sent through a sequence transformer, which results in a new set of vectors which have added context from neighbouring time periods. A small final block is then used to classify each epoch into one of the 5 sleep stages.

#### 2.2.6 AttnSleep

The AttnSleep algorithm [21] uses just one EEG channel (Fpz-Cz or C4-A1) as input to predict sleep stages. The model consists of 3 modules: 1) a multi-resolution CNN (MRCNN), designed to extract both low and high frequency features from a 30-second epoch, 2) a temporal context encoder (TCE), an multi-head attention block which encodes temporal information within the epoch, which is then fed to the 3) classification block. The model was trained and evaluated seperately on three dataset of healthy control subjects (ranging from 20 - 329 PSG recordings). Benchmarking against four prior state-of-the-art single-channel EEG models, AttnSleep achieved superior performance across wake, N2, N3, and REM.

**Table 4:**
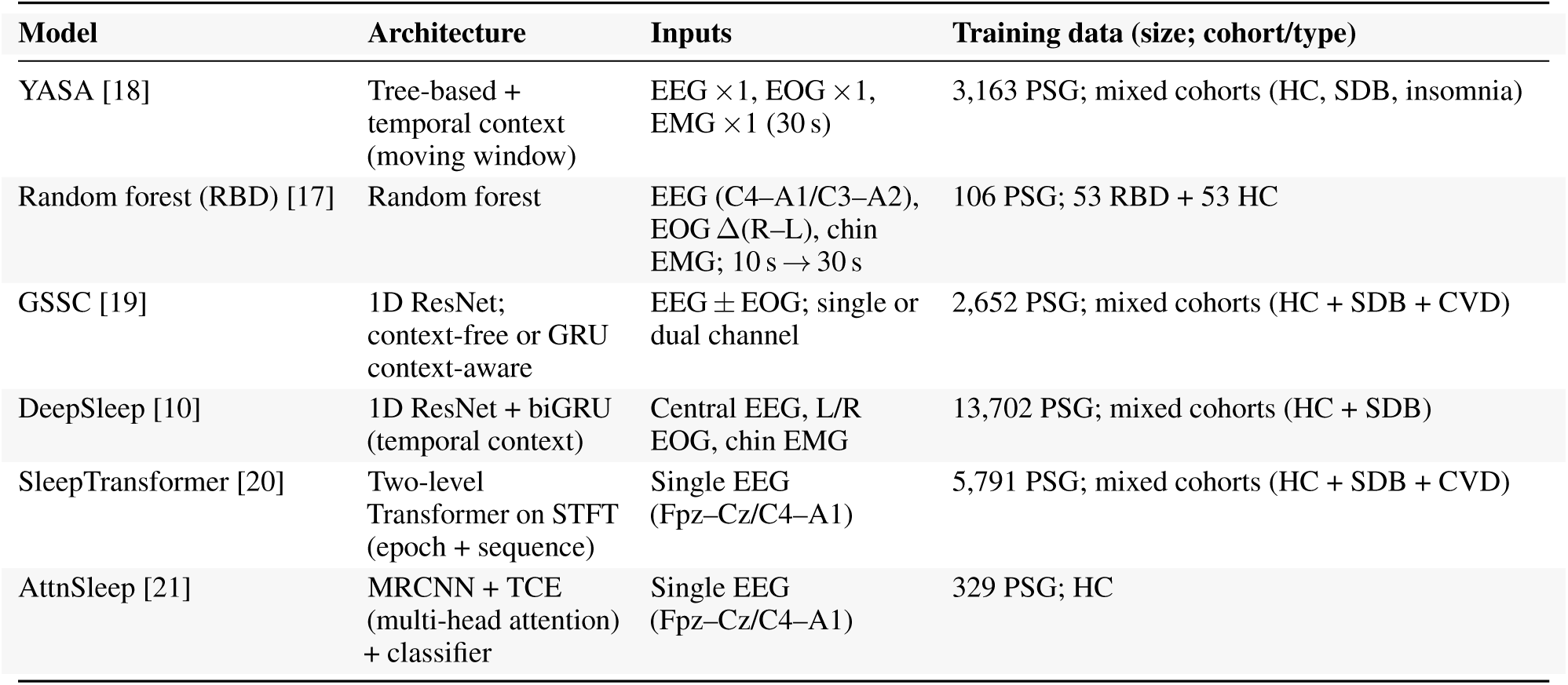
Summary of the six models compared in this work. CVD: cardiovascular disease. HC: healthy controls. SDB: sleep disordered breathing. STFT: short-time Fourier transform. GRU: gated recurrent unit. MRCNN: multi-resolution convolutional neural network. TCE: temporal context encoder.

| Model | Architecture | Inputs | Training data (size; cohort/type) |
| --- | --- | --- | --- |
| YASA [18] | Tree-based + temporal context (moving window) | EEG $\times 1$, EOG $\times 1$, EMG $\times 1$ (30 s) | 3,163 PSG; mixed cohorts (HC, SDB, insomnia) |
| Random forest (RBD) [17] | Random forest | EEG (C4-A1/C3-A2), EOG $\Delta$(R-L), chin EMG; 10 s → 30 s | 106 PSG; 53 RBD + 53 HC |
| GSSC [19] | 1D ResNet; context-free or GRU context-aware | EEG ± EOG; single or dual channel | 2,652 PSG; mixed cohorts (HC + SDB + CVD) |
| DeepSleep [10] | 1D ResNet + biGRU (temporal context) | Central EEG, L/R EOG, chin EMG | 13,702 PSG; mixed cohorts (HC + SDB) |
| SleepTransformer [20] | Two-level Transformer on STFT (epoch + sequence) | Single EEG (Fpz-Cz/C4-A1) | 5,791 PSG; mixed cohorts (HC + SDB + CVD) |
| AttnSleep [21] | MRCNN + TCE (multi-head attention) + classifier | Single EEG (Fpz-Cz/C4-A1) | 329 PSG; HC |

### 2.3 Preprocessing

As the clinical gold-standard and the portable PSG systems operate with independent internal clocks, temporal alignment was performed post hoc. Synchronisation between the PSG recordings was achieved by aligning recording start times. All analyses were restricted to the interval between lights off and lights on as annotated by the clinical sleep specialist in the vPSG recording. These annotations defined the ground-truth sleep period boundaries.

The same signal pre-processing procedures described in the original publications were applied to the portable PSG dataset prior to testing and fine-tuning. For YASA, signals were downsampled to 100 Hz, after which EEG channels were band-pass filtered between 0.4–30 Hz, Z-score normalised, and segmented into 30-second epochs. In the random forest RBD model, signals were first downsampled to 200 Hz prior to filtering. EEG and EOG channels were processed using a 500th-order FIR band-pass filter between 0.3–40 Hz, while EMG signals underwent 500th-order 50/60 Hz notch filtering followed by a 10–100 Hz FIR band-pass filter. No explicit normalisation procedure was reported. Features were extracted from 10-second windows and subsequently averaged to form 30-second epochs. Similarly, the GSSC model downsampled signals to 85.33 Hz before applying a 0.3–30 Hz zero-phase FIR band-pass filter. Z-score normalisation was performed independently for each channel prior to segmentation into 30-second epochs. DeepSleep preprocessing involved resampling signals to 128 Hz, followed by filtering of EEG and EOG channels between 0.5–35 Hz and high-pass filtering of EMG signals at 10 Hz using a zero-phase fourth-order IIR filter. Signals were then Z-score normalised and segmented into 30-second epochs, with additional temporal context incorporated through a bidirectional GRU architecture. In contrast, SleepTransformer did not employ conventional temporal-domain filtering or downsampling, instead operating on short-time Fourier transform (STFT) spectrogram representations of the signals. These spectrograms were Z-score normalised and processed as 30-second epochs. Finally, no explicit filtering or normalisation procedures were reported for AttnSleep, which operated directly on 100 Hz single-channel EEG data segmented into 30-second epochs.

### 2.4 Model training

#### Model comparison

The primary analysis was designed to compare the performance of the six sleep staging algorithms under a common held-out test. Recordings were split into training (65%), validation (15%), and test (20%) sets, stratified by diagnostic group (healthy control, RBD, and non-RBD sleep disorder patients) and acquisition site to ensure a balanced representation. Splitting was performed at the participant level, such that recordings from the same participant could not appear in more than one split. The validation set was used only for model selection during fine-tuning and was not used for the final model comparison, which was performed exclusively on the held-out test set.

Pretrained models were first applied directly to the held-out test set. The open-source models were then fine-tuned on the training set, with model selection based on validation performance, and subsequently evaluated on the same held-out test set. This primary held-out analysis was used to compare out-of-the-box and fine-tuned performance across all algorithms, both overall and by sleep stage. For the tree-based models (RF and YASA), no epoch-wise neural-network checkpoint selection was required. Therefore, the final retrained models were fitted on the training set and evaluated once on the held-out test set. All fine-tuning procedures used the hyperparameters reported in the original publications.

YASA is a tree-based gradient boosting classifier and does not use gradient-based optimisation. Class imbalance was handled via class weighting (Wake = 1.0, N1 = 2.2, N2 = 1.0, N3 = 1.2, REM = 1.4) as in the original publication [18]. No data augmentation was applied. The model is deterministic given fixed input features and therefore does not rely on batch size, learning rate scheduling, or early stopping. The model was trained for 500 boosting iterations. The RBD random forest is also a tree-based ensemble method and used 500 estimators.

GSSC was trained using the AdamW optimiser (*β*_1_ = 0.9, *β*_2_ = 0.999) with a learning rate of 3 *×* 10*^−^*^4^ and no learning rate scheduling. The *β* terms are hyperparameters that control how moving averages of gradients are computed. Training was performed for up to 20 epochs with all layers unfrozen. Weighted cross-entropy loss was used to address class imbalance (Wake = 1.0, N1 = 2.4, N2 = 1.0, N3 = 1.2, REM = 1.4). No data augmentation was reported. The model was trained for 20 epochs with all layers unfrozen. Model selection was based on accuracy on the validation set. Final performance was then reported on the held-out test set.

DeepSleep was trained using the Adam optimiser (*β*_1_ = 0.9, *β*_2_ = 0.999), with a learning rate of 1 *×* 10*^−^*^4^ and no learning rate scheduler. The authors did not apply class re-weighting or resampling to compensate for stage imbalance. The model was finetuned for 20 epochs with all layers unfrozen. Model selection was based on accuracy on the validation set. Final performance was then reported on the held-out test set.

SleepTransformer was trained using the Adam optimiser (*β*_1_ = 0.9, *β*_2_ = 0.999), *ɛ* = 1 *×* 10*^−^*^7^) with a learning rate of 1 *×* 10*^−^*^4^ and no learning rate scheduling. A batch size of 32 was used. No class weighting or resampling strategy was reported. No data augmentation was applied. All layers of the model were unfrozen during fine-tuning. Fine-tuning was performed for up to 100 epochs, with early stopping based on validation loss using a patience of 8 epochs. Model selection was based on accuracy on the validation set. Final performance was then reported on the held-out test set.

AttnSleep was trained using the Adam optimiser (*β*_1_ = 0.9, *β*_2_ = 0.999) with AMSGrad enabled, a learning rate of 1 *×* 10*^−^*^3^, and weight decay of 1 *×* 10*^−^*^3^. A batch size of 128 was used. Weighted cross-entropy loss was applied, with class weights computed from the training distribution. No data augmentation was applied. A fixed random seed (123) was used in the original implementation. Fine-tuning was performed for up to 100 epochs, with early stopping based on validation loss using a patience of 8 epochs. All layers were unfrozen during fine-tuning. Model selection was based on accuracy on the validation set. Final performance was then reported on the held-out test set.

#### Stability analysis of the best-performing model

To assess the robustness of the best-performing model to dataset partitioning, we performed an additional five-fold cross-validation analysis. The folds were divided at the participant level, ensuring that recordings from the same participant were not split across folds. Folds were stratified by diagnostic group and acquisition site to preserve the distribution of controls, patients without RBD, patients with RBD, and recording sites across folds.

For each cross-validation iteration, one fold was held out for evaluation and the remaining folds were used for model fine-tuning. The same preprocessing, model architecture, optimiser, loss function, class weighting, and hyperparameters as in the primary analysis were used. Performance was then computed on the held-out fold against the manual vPSG-derived sleep stage annotations.

For each cross-validation iteration, one fold was held out for evaluation and the remaining folds were used for model fine-tuning. The same preprocessing, model architecture, optimiser, loss function, class weighting, and hyperparameters as in the primary analysis were used. Performance was then computed on the held-out fold against the manual vPSG-derived sleep stage annotations.

For each fold, the selected model was fine-tuned using the same preprocessing, model architecture, optimiser, loss function, class weighting, and hyperparameters as in the primary analysis. The model checkpoint with the best validation performance was used to infer sleep stages for the corresponding validation fold. Performance was then computed against the manual vPSG-derived sleep stage annotations. This cross-validation analysis was used to assess whether overall performance was stable across dataset partitions and whether the stage-specific error profile varied across folds.

In addition to overall cross-validation metrics, per-stage cross-validation metrics were computed for Wake, N1, N2, N3, and REM. These per-stage metrics were calculated separately within each fold. The purpose of this analysis was to quantify whether some sleep stages showed greater fold-to-fold variability, as reflected by a larger standard deviation across folds, which would suggest less stable classification performance for those stages.

### 2.5 Evaluation metrics

#### Model comparison

We report the evaluation metrics for both the out-of-box application of the models and the fine-tuned models, on the portable PSG test set. Evaluation was performed at the epoch level. We report the overall accuracy, as well as the F1 score and Cohen’s *κ* overall and per sleep stage. Cohen’s *κ* is reported per sleep stage within each diagnostic group (controls, patients without RBD, and patients with RBD) to assess group-specific performance and the effects of fine-tuning across sleep stages.

The F1 score is the harmonic mean of recall (R) and precision (P), and is defined as:

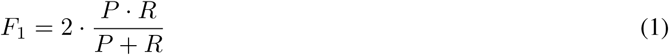

where the precision and recall are computed from the true positives (TP), false positives (FP), and false negatives (FN):

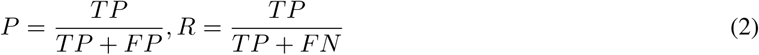

Cohen’s kappa (*κ*) is a measure of agreement between two raters, here the manual sleep staging and the output of the model. It is defined as:

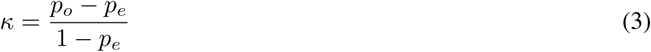

where *p_o_* is the observed agreement and *p_e_* is the expected agreement. The row-normalised confusion matrices of the results for each of the models are also reported.

#### 5-fold cross-validation

For the five-fold GSSC cross-validation analysis, the same overall metrics were computed on each held-out fold: accuracy, macro-F1, weighted-F1, and Cohen’s *κ*. These metrics were summarised as mean ± standard deviation across the five folds. Per-stage cross-validation performance was also computed for Wake, N1, N2, N3, and REM. For each sleep stage, F1 score and Cohen’s *κ* were calculated using a one-versus-rest approach within each held-out fold. Per-stage results were then summarised as mean ± standard deviation across folds.

## 3 Results

### Model comparison

Sleep staging data were available for the 59 subjects across three clinical sites, comprising 76,940 thirty-second epochs in total (Table 5). Epochs across the three diagnostic groups is also reported: healthy controls (n=14), patients without RBD (n=38), and patients with RBD (n=24). The stage distribution was consistent with typical clinical populations, with N2 being the most prevalent stage across all groups (35.5–39.2%), followed by Wake (21.6–24.7%), N3 (14.1–16.0%), REM (12.8–13.6%), and N1 (8.8–12.2%). The RBD group exhibited a slightly higher proportion of Wake (24.7% vs 21.6–22.9%) and N1 (12.2% vs 8.8–11.3%) epochs, alongside a lower proportion of N2 (35.5% vs 38.9–39.2%). Controls had the lowest proportion of N1 (8.8%), consistent with fewer arousals and more consolidated sleep. The overall class imbalance, particularly the low prevalence of N1, is relevant to interpreting the per-stage classification performance reported below.

The overall performance of each of the sleep staging models using the pretrained model and the model fine-tuned on our in-clinic dataset is shown in Table 6. The highest performance for both model settings is indicated in bold. The fine-tuned models consistently outperformed the pre-trained models across all metrics. The highest performance was achieved by the fine-tuned GSSC model, with a *κ* of 0.58.

**Table 5:**
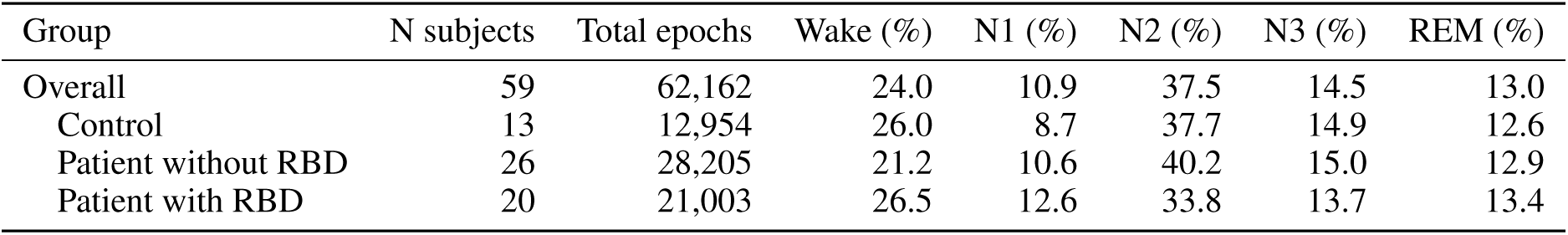
Sleep stage percentage distribution across diagnostic groups.

| Group | N subjects | Total epochs | Wake (%) | N1 (%) | N2 (%) | N3 (%) | REM (%) |
| --- | --- | --- | --- | --- | --- | --- | --- |
| Overall | 59 | 62,162 | 24.0 | 10.9 | 37.5 | 14.5 | 13.0 |
| Control | 13 | 12,954 | 26.0 | 8.7 | 37.7 | 14.9 | 12.6 |
| Patient without RBD | 26 | 28,205 | 21.2 | 10.6 | 40.2 | 15.0 | 12.9 |
| Patient with RBD | 20 | 21,003 | 26.5 | 12.6 | 33.8 | 13.7 | 13.4 |

**Table 6:** Classification performance of different models and training settings. Highest performance for fine-tuned and pre-trained settings indicated in bold.

| Model | Setting | Accuracy | F1 macro | F1 weighted | Cohen's $\kappa$ |
| --- | --- | --- | --- | --- | --- |
| AttnSleep | Pretrained | 0.35 | 0.34 | 0.34 | 0.21 |
| AttnSleep | Finetuned | 0.44 | 0.43 | 0.45 | 0.29 |
| DeepSleep | Pretrained | 0.44 | 0.36 | 0.41 | 0.25 |
| DeepSleep | Finetuned | 0.52 | 0.48 | 0.50 | 0.36 |
| GSSC | Pretrained | **0.66** | **0.61** | **0.63** | **0.54** |
| GSSC | Finetuned | **0.68** | **0.65** | **0.67** | **0.58** |
| SleepTransformer | Pretrained | 0.45 | 0.36 | 0.40 | 0.28 |
| SleepTransformer | Finetuned | 0.63 | 0.56 | 0.60 | 0.50 |
| RF | Pretrained | 0.45 | 0.40 | 0.43 | 0.27 |
| RF | Finetuned | 0.47 | 0.41 | 0.44 | 0.29 |
| YASA | Pretrained | 0.56 | 0.50 | 0.54 | 0.41 |
| YASA | Finetuned | 0.61 | 0.54 | 0.58 | 0.46 |

Figure 1 shows the cohen’s *κ* per sleep stage for each of the pretrained and fine-tuned models. This is shown overall and by subject group (control, patient without RBD and patient with RBD). Across all subjects, GSSC attains the highest *κ* in wake, N1, N2, and REM for the pretrained and finetuned model.

**Figure 1:**
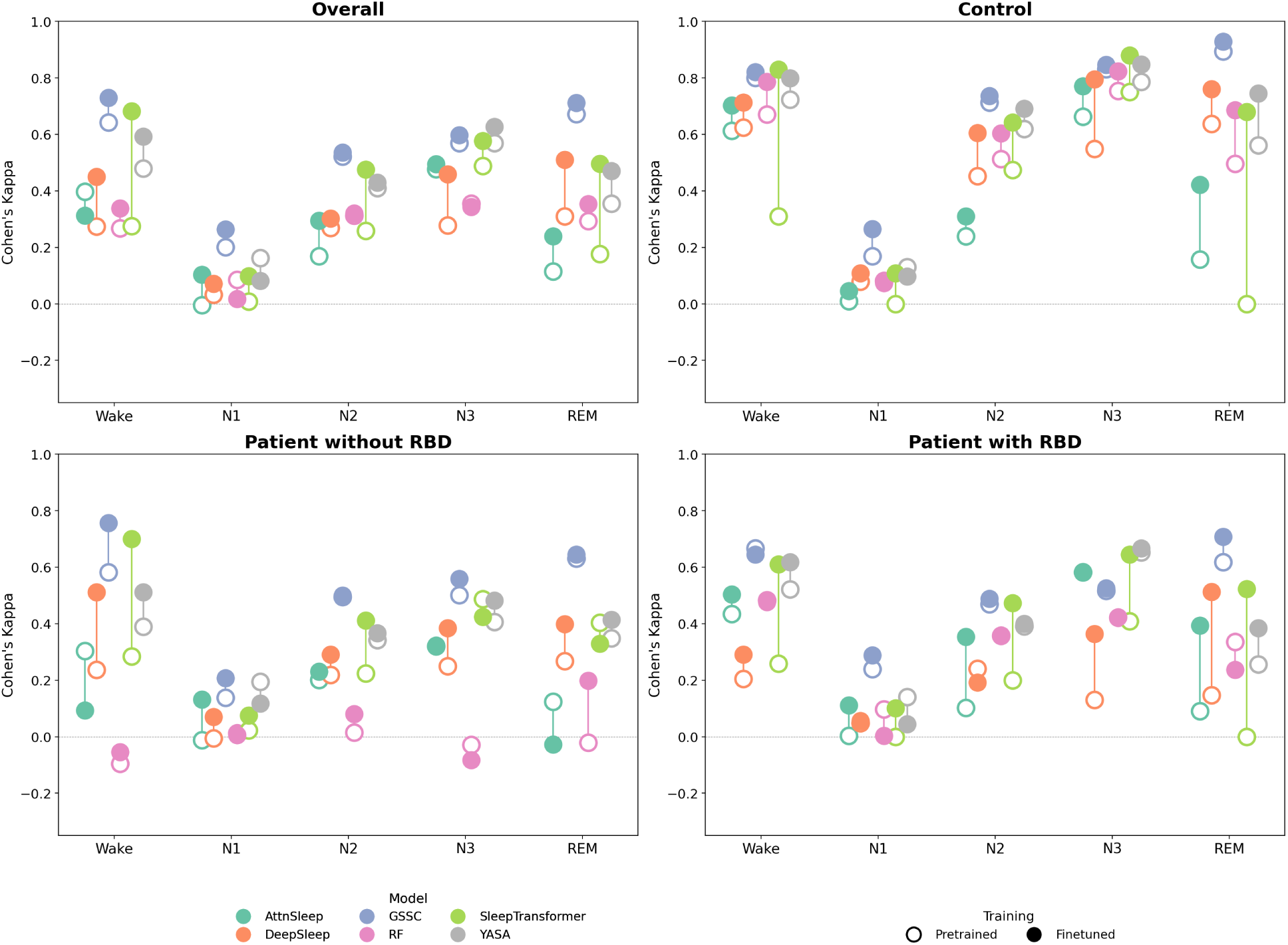
Cohen’s *κ* across sleep stages for pretrained and fine-tuned models applied to the test dataset overall and by subject diagnosis group.

### 5-fold cross-validation

Following the primary held-out model comparison, the best performing model, GSSC, was evaluated using five-fold cross-validation. For each cross-validation iteration, the model was fine-tuned on the corresponding training folds and evaluated on the held-out fold. Table 7 shows the mean fold-level performance together with the mean within-fold standard deviation for each overall metric.

**Table 7:** Five-fold cross-validation performance of the fine-tuned GSSC model. Metrics were computed on the held-out fold in each cross-validation iteration and are reported as the mean fold-level performance *±* the mean within-fold standard deviation.

| Metric | Mean ± SD |
| --- | --- |
| Accuracy | $0.708 \pm 0.171$ |
| Macro-F1 | $0.618 \pm 0.156$ |
| Weighted-F1 | $0.704 \pm 0.177$ |
| Cohen’s $\kappa$ | $0.597 \pm 0.219$ |

Per-stage performance was computed for each validation fold using a one-versus-rest formulation for each sleep stage. Table 8 shows the mean and standard deviation across folds for the per-stage F1 score and Cohen’s *κ*.

**Table 8:**
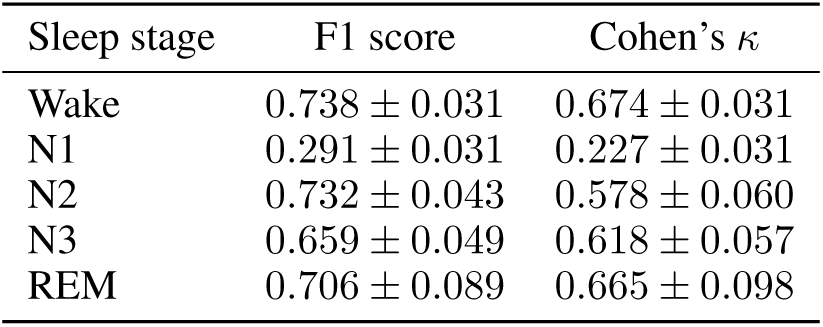
Per-stage five-fold cross-validation performance of the fine-tuned GSSC model. Per-stage F1 score and Cohen’s *κ* were computed using a one-versus-rest formulation and are reported as mean *±* standard deviation across folds.

| Sleep stage | F1 score | Cohen’s $\kappa$ |
| --- | --- | --- |
| Wake | $0.738 \pm 0.031$ | $0.674 \pm 0.031$ |
| N1 | $0.291 \pm 0.031$ | $0.227 \pm 0.031$ |
| N2 | $0.732 \pm 0.043$ | $0.578 \pm 0.060$ |
| N3 | $0.659 \pm 0.049$ | $0.618 \pm 0.057$ |
| REM | $0.706 \pm 0.089$ | $0.665 \pm 0.098$ |

## 4 Discussion

### 4.1 Overall performance

This study presents the first comparison of multiple open-source sleep staging algorithms on a minimal portable PSG set-up in patients with and without RBD. Six open-source algorithms were applied to our SleepWearables portable PSG dataset. The overall performance of the open-source algorithms varied; with Cohen’s *κ* values ranging between 0.21 and 0.54, when applied out-of-box. Fine-tuning the open-source algorithms on the portable PSG data improved performance across all models, however the degree of improvement varied. DeepSleep and SleepTransformer had the largest gains in performance after fine-tuning (*κ* 0.25 to 0.36, and 0.28 to 0.50, respectively). The fine-tuned GSSC achieved the highest agreement (*κ* = 0.58).

An improvement in performance following fine-tuning is expected, as the models can capture population-specific features. Both GSSC and YASA demonstrated moderate agreement (*κ >* 0.40), with the fine-tuned GSSC approaching *κ* = 0.60, indicative of substantial agreement [27]. Models that combine feature extraction with temporal context (e.g., GSSC’s ResNet+GRU and YASA’s feature-based moving window) can benefit from both within-epoch feature learning and contextual smoothing, improving generalisability to a reduced-channel portable montage. Whilst DeepSleep and the SleepTransformer also incorporate context from neighbouring epochs, these models are more complex. The DeepSleep model uses a deeper ResNet architecture than GSSC in order to maximise the representational capacity of the resulting features. However, increased model depth and capacity are well-established factors that increase the risk of overfitting to the training data distribution, particularly in settings with limited or homogeneous data [28]. Similarly, the SleepTransformer uses a transformer-based architecture to extract both epoch and sequence level features. Transformers are known to require substantially more data to fine-tune to a new data distribution [29].

AttnSleep attained a *κ* of 0.29 after fine-tuning, remaining well below the best-performing algorithms. This may be attributed to the model having been pretrained on a single healthy control dataset. The extracted features might not be representative of sleep architecture in clinical populations. As with SleepTransformer, AttnSleep’s transformer block may require more training data to generalise effectively to our clinical distribution. Whilst the RF model was trained on subjects with RBD in addition the healthy controls, it was trained on a small cohort of only 106 PSGs from 106 subjects. Training the RF on our clinical dataset improved performance marginally (*κ* increased from 0.27 to 0.29), suggesting that limited training data or model architecture remain a key bottleneck.

Fine-tuned models (particularly GSSC) reached moderate agreement and may support screening and reducing manual review time. However, for diagnostic decisions, e.g. confirming RBD, current performance (especially for REM detection) requires review in comparison to the inter-rater variability in a real-life clinical population including patients with RBD. This is analogous to the use of home sleep apnea testing (HSAT) in sleep apnea, which is suitable for initial assessment but often requires follow-up with full polysomnography when results are negative, inconclusive, or when clinical suspicion remains high [30]. Notably, a larger pretraining dataset or inclusion of multiple cohorts did not necessarily yield better performance on our data, challenging the commonly held assumption that increasing pretraining scale alone improves generalisation in automatic sleep staging.

### 4.2 Per group and sleep stage performance

The stage-wise Cohen’s *κ* values provide a more granular view of model behaviour than overall agreement. Because per-stage *κ* is effectively a one-versus-rest measure, it is influenced by stage distributions. This is particularly relevant for N1, which shows consistently low *κ* across all models and subgroups. The poor N1 agreement likely reflects a combination of low prevalence, transitional physiology between Wake and N2, and intrinsic inter-scorer variability at stage boundaries [2, 31]. Consequently, low N1 *κ* should not be interpreted purely as algorithmic results, but as a limitation of the staging problem that is amplified in reduced-channel recordings.

Across all subgroups, a consistent hierarchy of stage difficulty is observable. N3 demonstrates the highest and most stable agreement, particularly in controls, which is expected as slow-wave morphology remains detectable even on reduced montages. N2 generally reaches moderate agreement and improves after fine-tuning in models that were initially mismatched to the portable montage. Wake achieves high *κ* in controls and moderate agreement in patients, but becomes more variable in clinical cohorts, likely due to sleep fragmentation and increased ambiguity at Wake–N1 and Wake–REM transitions [31]. REM is the most model- and subgroup-dependent stage, with large improvements after fine-tuning in several architectures but persistent underperformance in some models. N1 remains the least well-classified stage in all analyses.

In the control cohort, most models achieve high agreement for Wake, N2, N3, and REM after fine-tuning, and some perform well before fine tuning. The largest improvements are observed in models that appear most sensitive to domain shift. For example, the SleepTransformer shows substantial gains in Wake and REM after fine-tuning, indicating that its transformer-based sequence modelling can perform strongly on the reduced montage once adapted, but is highly dependent on in-domain training. DeepSleep shows a similar pattern with an architecture benefiting from adaptation to the portable clinical distribution. Despite these improvements, N1 agreement remains low even in controls, reinforcing that this stage represents an inherent limitation rather than a pathology-specific issue.

In patients without RBD, performance is generally lower and more heterogeneous. GSSC remains among the strongest performers across Wake, N2, and N3 after fine-tuning, suggesting robust generalisability. SleepTransformer benefits substantially from adaptation, particularly in Wake and deeper stages. In contrast, AttnSleep demonstrates instability in this subgroup, with a decrase in performance in Wake and REM agreement after fine-tuning, including near-zero or negative *κ* in some stages. This pattern is consistent with limited generalisation following fine-tuning on a small dataset. The RF baseline performs poorly in this cohort, with negative or near-zero *κ* in several stages and only limited improvement after retraining, suggesting that fixed feature representations may be insufficient to capture the heterogeneity of clinical sleep without more complex modelling.

Challenges in REM and Wake staging are also observed in the RBD group. Pre-trained model agreement is lower, but fine-tuning leads to improvements in several deep learning models. SleepTransformer and DeepSleep show gains in REM agreement after adaptation, indicating that complex models can learn cohort-specific REM characteristics under altered EMG tone and increased movement. GSSC achieves the highest overall performance in this subgroup after fine-tuning, with moderate agreement in REM and stable performance in N2 and N3, suggesting robustness to montage reduction and pathological sleep architecture. YASA improves in Wake and N3 but remains weaker in REM, consistent with the limitations of engineered feature sets under pathological conditions. The RF model shows inconsistent behaviour and decreased REM agreement after retraining in RBD, reinforcing concerns that its feature representation and model capacity are insufficient for this subgroup and PSG montage.

The magnitude of the pretraining-to-fine-tuning improvements provides insight into domain shift and model complexity. Small performance differences combined with strong absolute *κ*, as seen in GSSC across several stages, indicate transferability and reduced sensitivity to distributional mismatch. Large differences from poor to strong performance, particularly in SleepTransformer and DeepSleep indicate reliance on in-domain adaptation. In contrast, inconsistent or negative differences, especially in AttnSleep and the RF model for certain subgroups and stages, suggest instability or overfitting under limited data conditions and highlight the need for more data and conservative fine-tuning strategies.

From a clinical perspective, N3 appears to be the most reliably automated stage across models after fine-tuning, supporting the use of deep sleep estimates in derived sleep metrics. N2 achieves moderate reliability and may be adequate sleep architecture summaries. Wake and REM, which are required for measures such as sleep efficiency and REM proportion and are particularly relevant in RBD, show meaningful improvement with fine-tuning in the best-performing models but should still require expert review in diagnostic contexts. N1 detection remains the principal challenge across all subgroups, limiting the reliability of sleep onset metrics and fine-grained transition analyses.

Several limitations should also be acknowledged. A total of 17 PSG recordings (22%) were excluded from analysis. Poor chin EMG signal quality accounted for 11 of these exclusions (65%), likely reflecting the increased susceptibility of the bulkier chin EMG electrode to signal loss during overnight recording. Thus, the test set only included 12 patients and PSGs. Whilst there have not been any studies on signal quality or electrode detachment in the specific portable-PSG kit used in this study, signal acquisition failure is a common issue in portable-PSG due to electrode misplacement or detachment during the overnight recording [32]. This is likely exacerbated in patients with RBD or other sleep disorders which result in increased movement during sleep. Given the small test set, especially within subgroups, these stage-specific findings should be interpreted cautiously. Moreover, as staging agreement in clinical settings can vary substantially between human reviewers, the present study cannot determine how the best automated performance compares to the agreement achievable between expert scorers in these datasets [33]. The per-stage analysis reinforces the conclusion that fine-tuning mitigates domain shift effects, that model architecture influences robustness under reduced-channel clinical conditions, and that REM and N1 remain the key bottlenecks for clinical-grade automatic sleep staging.

### 4.3 5-fold cross-validation

The five-fold cross-validation analysis showed that the fine-tuned GSSC model achieved similar overall performance to that observed in the primary held-out test analysis. Across folds, GSSC reached an accuracy of 0.708, weighted-F1 of 0.704, macro-F1 of 0.618, and Cohen’s *κ* of 0.597. These values are close to the held-out test performance of the fine-tuned GSSC model, supporting that its performance was not due to favourable single train-validation-test split. The cross-validation results suggest that GSSC generalised relatively consistently across different partitions of dataset.

Weighted-F1 was substantially higher than macro-F1, indicating that performance was stronger for the more prevalent sleep stages and lower for less frequent or more ambiguous stages. This is expected in sleep staging datasets, where N2 and Wake are typically prevalent in the epoch distribution, while N1 is less common and more difficult to classify. Although the overall accuracy and weighted-F1 suggest good aggregate performance, the lower macro-F1 indicates that performance was not uniform across all stages. This supports the need to interpret overall metrics together with stage-specific performance rather than relying on accuracy alone.

The mean within-fold standard deviations were relatively large for all overall metrics, particularly for Cohen’s *κ* (*±*0.219), weighted-F1 (*±*0.177), and accuracy (*±*0.171). This indicates variability in performance between recordings within the same validation fold. Such variability is expected in this dataset, given the heterogeneity of the clinical cohort, differences in sleep architecture, recording quality, site of acquisition, and diagnostic groups. It also shows that automatic sleep staging performance can vary at the individual level, even when average fold-level performance appears stable.

The low variability of the fold-level mean performance indicates that the overall GSSC result is robust to dataset partitioning. Moreover, the larger within-fold standard deviations suggests that individual-recording performance remains variable. This distinction is clinically meaningful: while the model shows stable average performance across cross-validation folds, its reliability for a given individual recording may still depend on recording quality, sleep-stage composition, and clinical phenotype. The cross-validation results strengthen the confidence in GSSC as the most robust model in this comparison, but also reinforce that automatic staging should be used as a supportive tool rather than as a fully autonomous replacement for expert scoring in clinical populations.

The per-stage five-fold cross-validation analysis showed that GSSC performance varied substantially across sleep stages. Wake, N2, N3, and REM achieved substantially higher F1 scores and Cohen’s *κ* values than N1, indicating that the stage-wise pattern observed in the primary held-out analysis was also present across cross-validation folds. N1 had the lowest performance, with an F1 score of 0.291 *±* 0.031 and Cohen’s *κ* of 0.227 *±* 0.031. The low mean values indicate that N1 remained the least accurately classified stage, while the small standard deviations show that this poor performance was consistent across folds rather than being driven by a single unfavourable partition.

Wake showed the highest F1 score among the sleep stages (0.738 *±* 0.031), together with a high Cohen’s *κ* (0.674 *±* 0.031). The standard deviations suggest that Wake classification was stable across folds. N2 also showed better performance, with an F1 score of 0.732 *±* 0.043 and Cohen’s *κ* of 0.578 *±* 0.060. This is consistent with N2 being one of the most prevalent stages in the dataset, which likely provides the model with more examples during training and contributes to more stable fold-level performance.

N3 showed moderate-to-high agreement, with an F1 score of 0.659 *±* 0.049 and Cohen’s *κ* of 0.618 *±* 0.057. Although its F1 score was lower than Wake and N2, its Cohen’s *κ* was comparable to N2, suggesting that GSSC captured N3 adequately across folds. This is likely because N3 is characterised by distinctive slow-wave activity, which remains detectable even with a reduced portable montage. The moderate standard deviations indicate some fold-to-fold variability, but less than that observed for REM.

REM achieved a relatively high mean F1 score (0.706 *±* 0.089) and Cohen’s *κ* (0.665 *±* 0.098), but showed the largest standard deviation across folds. This suggests that REM classification could be strong in some folds but less stable across dataset partitions. This variability is clinically relevant in the context of RBD, where REM sleep is central to diagnosis and where abnormal motor activity, altered REM physiology, and movement artefacts may make REM classification more dependent on the specific subjects included in each fold. Although the mean REM performance was encouraging, the higher fold-to-fold variability suggests that REM classification remains less robust than Wake or N2.

Overall, the per-stage cross-validation analysis supports the interpretation that GSSC performance is stage-dependent. N1 was consistently the weakest stage, but not the most variable, indicating a systematic limitation rather than fold-specific instability. REM had relatively high average performance but the greatest variability, suggesting that it is more sensitive to dataset composition. Wake and N2 showed the most stable performance across folds, while N3 remained moderately stable with good agreement. These findings reinforce the need to report stage-specific metrics in addition to overall performance, particularly in clinical cohorts where REM and N1 errors may have diagnostic relevance.

## 5 Conclusion

This study compared multiple open-source sleep staging algorithms on a reduced-channel portable PSG setup in controls and patients with and without RBD. Fine-tuning on in-clinic data consistently improved performance across models, underscoring the importance of domain adaptation when transferring pretrained models to new PSG montages and clinical populations. GSSC demonstrated the most robust overall performance and the strongest generalisability across subgroups. More complex architectures, including transformer-based models, showed the largest improvements after fine-tuning, indicating higher capacity for domain adaptation. In contrast, models relying on fixed feature representations or limited pretraining were less stable and showed smaller gains. Stage-wise analysis confirmed N3 as the most reliably detected stage and N1 as the most challenging across all subgroups. REM classification improved with fine-tuning in several models but remained variable, particularly in RBD. Although fine-tuned models achieved moderate agreement and may support clinical workflows, performance, especially for REM and N1, remains insufficient for fully automated clinical use.

## Data Availability

Access to the Oxford Parkinson's Disease Centre (OPDC) SleepWearables PSG dataset used in this study is available through a formal submission to the Data Access Committee who will review this and either support, decline, or request further information. See https://www.dpag.ox.ac.uk/opdc/research/external-collaborations for more information.

